# Bias amplification of unobserved confounding in pharmacoepidemiological studies using indication-based sampling: there is no free lunch in restricting the sample to those with a particular drug-indication

**DOI:** 10.1101/2022.02.07.22270609

**Authors:** Viktor H. Ahlqvist, Paul Madley-Dowd, Amanda Ly, Jessica Rast, Michael Lundberg, Egill Jónsson-Bachmann, Daniel Berglind, Dheeraj Rai, Cecilia Magnusson, Brian K. Lee

## Abstract

Estimating causal effects in observational pharmacoepidemiology is a challenging task, as it is often plagued by confounding by indication. Restricting the sample to those with an indication for drug use is a commonly performed procedure; *indication-based sampling* ensures that the exposed and unexposed are exchangeable on the indication - limiting the potential for confounding by indication. However, indication-based sampling has received little scrutiny, despite the hazards of exposure-related covariate control.

Using causal diagrams, simulations, and empirical examples, we demonstrate that indication-based sampling in the presence of unobserved confounding can give rise to bias amplification, a self-inflicted phenomenon where one inflates pre-existing bias through inappropriate covariate control. Additionally, we show that indication-based sampling generally leads to a greater net bias than alternative approaches, such as regression adjustment. Finally, we expand on how bias amplification should be reasoned about when distinct clinically relevant effects on the outcome among those with an indication (effect-heterogeneity) exist.

We conclude that studies using indication-based sampling should have robust justification - and that it should by no means be considered a “free lunch” to adopt such approaches. As such, we suggest that future observational studies stay wary of bias amplification when considering drug indications.

## Introduction

Studies of the effectiveness and safety of medications are often biased by confounding by indication when utilizing observational data, sometimes referred to as *a most stubborn bias* [1]. Specifically, the indication for drug use may influence the outcome of interest, independently of the exposure, which induces confounding in the estimate of the drug-outcome relationship. This is less of an issue when the outcome is unintended (an unexpected consequence or benefit), as is the case in studies of drug safety and repurposing. When the indication for treatment is inherently linked to the outcome, as is the case when the outcome is intended, this confounding may instead be paramount. Hence new methods, such as those considering active comparators (i.e., comparing the drug of interest to another commonly used medication with a known effect for the same indication) [2, 3], circumvent confounding by indication by ensuring that everyone in the study shares the indication. Other frameworks, such as target trial emulation, are useful when reasoning about various selection mechanisms [4, 5]. Yet, some of these methods may be problematic. We here introduce how methods that rely on *indication-based sampling* may indeed exaggerate, rather than alleviate, bias in observational studies of drug effects.

Indication-based sampling is a procedure commonly performed in pharmacoepidemiology where one selects a cohort of individuals with an indication for drug use, and sometimes the absence of contraindication, either from a larger data frame (e.g., electronic health/medical records data) [6-9] or by enrolling participants into a primary cohort [10]. Such an approach is sometimes colloquially referred to as *restriction*, here we coin the term indication-based sampling to emphasise its orientation around drug indication. The motivation for performing indication-based sampling is often to make individuals exposed and unexposed to the drug under study near-identical regarding the indication (exchangeable), thereby reducing the potential for confounding by indication.

As such, using indication-based sampling, researchers aim to ensure perfect balance on a key determinant of drug use. We must then ask ourselves, however – why are some individuals with the indication using the drug? And why are some not? There must be factor(s) that drive this difference, assuming it is not purely stochastic. If the factor(s) has some independent effect on the outcome (i.e., meet the criteria of a classic confounder), the researchers will have unwittingly amplified its potential to bias the drug-outcome association as the exposed and unexposed are more likely to be discordant on it. In other words, by removing the information in the exposure that is explained by the indication, we amplify the influence of other, potentially unknown factors, that influence the exposure and potentially the outcome. Such a phenomenon is known as bias amplification [11]. Importantly, this phenomenon occurs even if the indication for drug use is a classic confounder [11]. That is, the bias amplification potential of selection on the indication is independent of the confounding originating from the indication. In fact, this amplified bias may distort associations between treatment and outcome to a larger extent than confounding by indication itself. This is particularly true when the unobserved confounding of the treatment and the outcome outweighs the confounding from indication, as one might expect in studies where the outcome is *unintended*.

Yet, we are unaware of any previous description of how bias amplification arises under indication-based sampling. There are previous descriptions in the econometric literature of bias amplification in propensity score analysis [12, 13], and detailed descriptions of the phenomena in the causal inference literature [11, 14-16]. Furthermore, it has, to our knowledge, not been demonstrated that indication-based sampling may amplify bias to a *greater* degree than when conditioning on drug indication. As indication-based sampling is common in pharmacoepidemiology and health technology assessments, elucidation of the potential for bias amplification through this method of design is important for future studies.

Here, we provide a description of bias amplification under indication-based sampling, which we highlight using simulations of varying levels of confounding and applied examples from pharmacoepidemiology, overall and in relation to effect-heterogeneity. To guide applied analysts, we contrast the amount of bias amplification under indication-based sampling to that of standard regression adjustment in an unrestricted sample. Ultimately, we showcase how and when indication-based sampling may distort measures of treatment effects over and beyond the *most stubborn bias of confounding by indication* [1] and thus when such sampling should be avoided counter to current practice and in spite of recent enthusiasm for active-comparators and target trial emulation in observational research.

### Informal description of bias amplification

The bias from indication-based sampling can be appreciated if one conceptualizes indication as an instrumental variable (i.e., a factor that has a causal effect on the exposure and that does not influence the outcome of interest except via the exposure). Specifically, it is possible to conceptualize drug indication as an instrumental variable (IV) of the exposure (Figure 1A), regardless of whether it is a perfect IV or not. It has been demonstrated in theoretical work [11-16] and reiterated in epidemiological literature [17-20], that conditioning on IVs induces bias amplification of unobserved confounders (U). That is, the bias from U on the exposure-outcome association becomes amplified. It has been further shown that this is true even when the IV has some non-zero effect on the outcome of interest (a so-called ‘Near-IV’[17]), making them indistinguishable from informal epidemiological definitions of confounders (Figure 1B).

**Figure 1.**
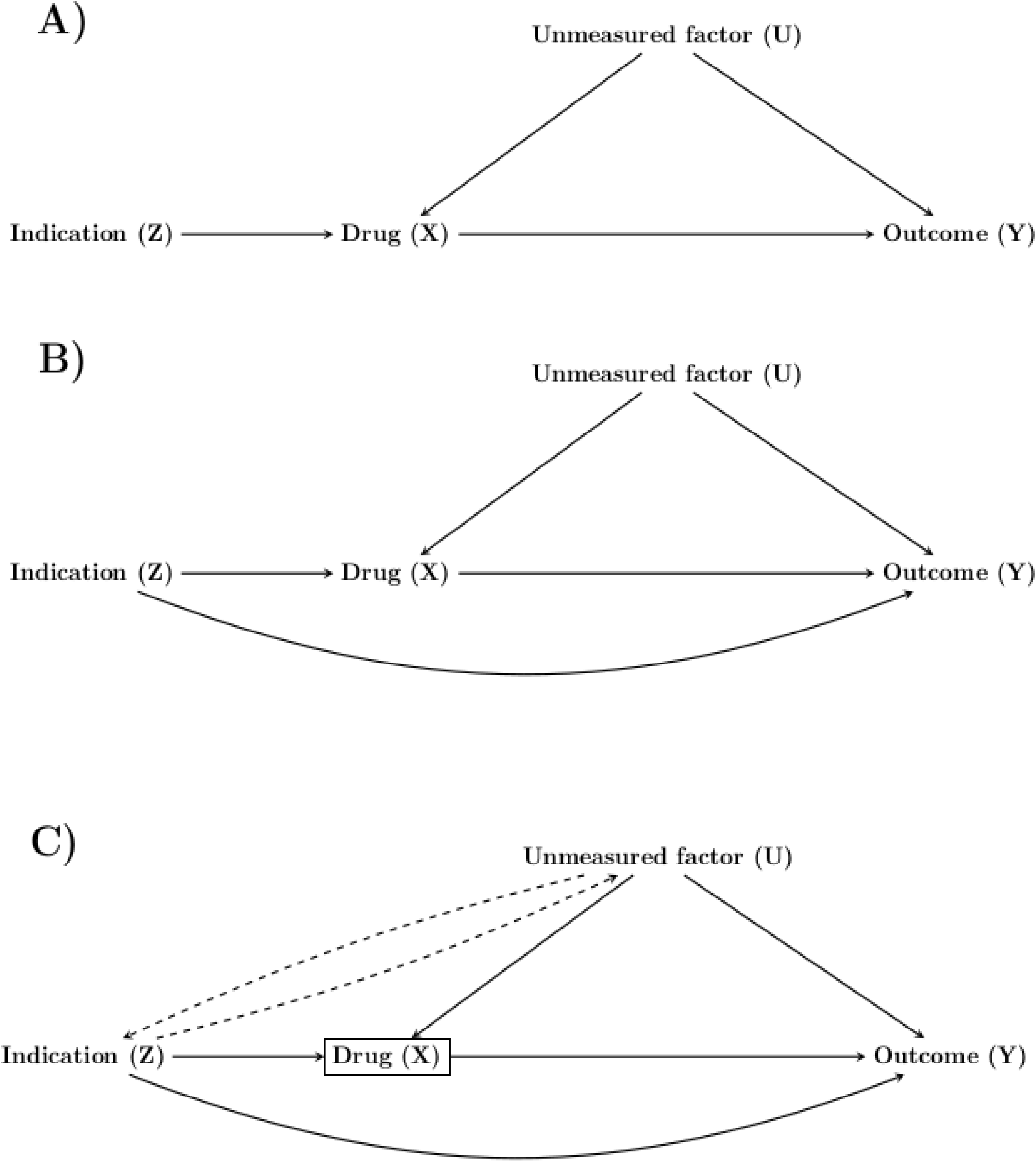
Directed acyclic graphs showing drug indication as a perfect IV (A), as a near-IV (B), and as a near-IV with induced correlation with the unmeasured factor due to collider stratification on drug use (C).

Pearl [11] provides an intuitive description of bias amplification under the same directed acyclic graph as in Figure 1A. According to Pearl, if we allow Z (the indication) to vary freely, Z will explain some of the differences in X (drug use). However, if we constrain Z=z, a larger share of the variation in X must be due to U (an unmeasured confounder). As such, we are now under maximized confounding from U. In other words, part of the difference in drug use will be due to indication and some will be due to the unmeasured confounder. If we remove the influence of the indication through indication-based sampling, we increase the share of variation in drug use that is explained by the unobserved confounder – maximizing its potential to bias our estimate. Yielding an answer to our initial question: why are some individuals using the drug? Simply because they differentially experience the confounder under indication-based sampling.

It is also possible to reason about bias amplification from a collider perspective [21], where colliders can be colloquially defined as factors that are influenced by at least two other factors in the causal system. Specifically, drug use (X) is a collider because of the effect of the indication (Z) and the unmeasured confounder (U) on drug use (Figure 1C). When attempting to estimate the causal effect of drug use on the outcome we condition on drug use (e.g., by fitting a regression model). However, drug use is a collider, and we thus induce a correlation between Z and U. This induced bias is smaller in magnitude and the opposite direction of the original confounding from U. When conditioning Z=z we are removing such offsetting collider bias. This, in turn, leads to a net bias which is greater than had we not conditioned on Z. We refer the readers to Wyss et al. [21] for an elegant description of how bias amplification arises due to offsetting effects under collider stratification. Importantly, this offsetting collider mechanism is invariant of Z’s possible influence on Y. As such, this bias mechanism is present independently of whether or not Z qualifies as a classic confounder, leading to the conclusion that the offsetting collider bias should be weighed against the possible confounding introduced by Z [11].

To reiterate, conditioning on a ‘near-IV’ that is associated with the outcome through a path not mediated by the exposure will result in amplification of other factors (here, the ‘near-IV’ is indistinguishable from a confounder) [17]. Under scenarios that we outline in this work, controlling for the IV will result in a greater bias in the exposure-outcome association than had we not controlled for the IV (despite the IV being a classic confounder). Thereby reinforcing the somewhat counterintuitive notion that controlling for a factor that would typically be labelled a confounder might increase net bias. Thus, while it is true that indication-based sampling can eliminate confounding by indication, it is not necessarily true that net bias will be less than when disregarding drug indication. To the best of our knowledge, this perplexing phenomenon has not yet been recognized in pharmacoepidemiology -despite its implications for causal inference and health technology assessments.

### A hypothetical real-world example where indication-based sampling suffers the greatest net bias: Statins and lung cancer

Let us consider a hypothetical real-world example of the causal structure in Figure 2. We want to study the effect of statin use (X) on incident lung cancer (Y), as others have done [22], in Swedish health registries. We have a clinical interest in the patient population with familial hypercholesterolemia (Z), an autosomal dominantly inherited disorder resulting in elevated low-density lipoprotein cholesterol (LDL-C) for which statins are indicated. Therefore, we perform indication-based sampling and only study those with a recorded diagnosis of familial hypercholesterolemia (Z=1). Unfortunately, as is often the case when using electronic health databases, we have no information on smoking (U) – a strong confounder in our relation-ship of interest.

**Figure 2.**
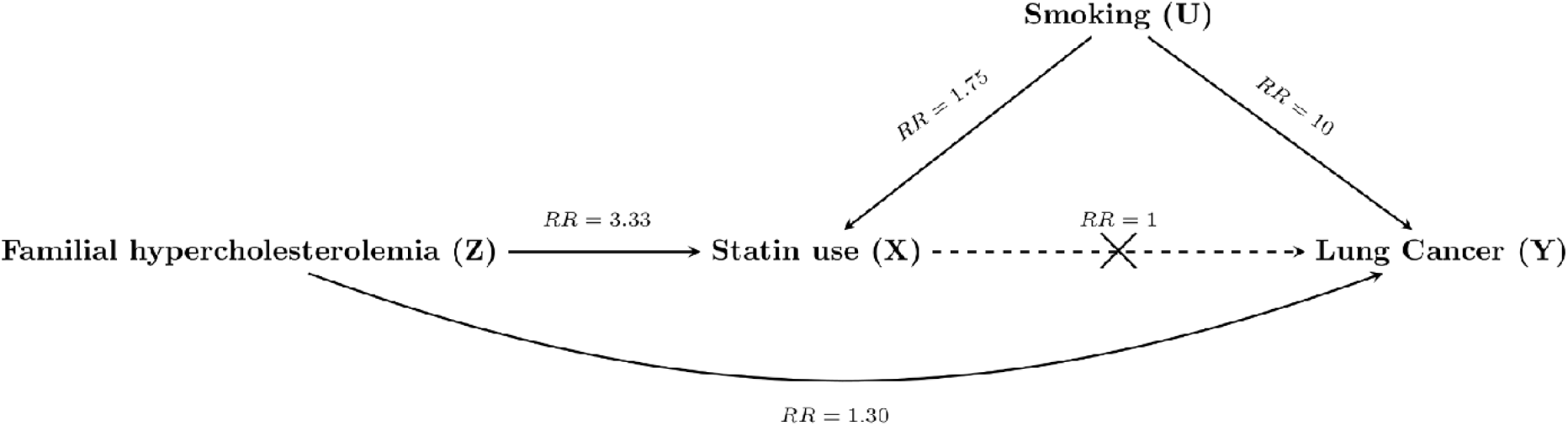
Directed acyclic graph showing a hypothetical real-world example of the relationship between familial hypercholesterolemia (Z), statin use (X), smoking (U), and lung cancer (Y), and the magnitude of their relationships (RR, relative risk), where the causal effect of statin use on lung cancer is of interest (but there is no such effect, RR=1).

Given that we are performing our study in Sweden, we expect the baseline risk of lung cancer to be ∼0.1%, statin use to have a prevalence of ∼15% and smoking to have a prevalence of ∼10%. Familial hypercholesterolemia has a prevalence of ∼1:200 and statin use has been reported to be ∼50% in clinical cohorts of this population (therefore, RR: Z→X=3.33) [23]. For simplicity, we will assume that those with familial hypercholesterolemia smoke at the same rate as the general population of Sweden (i.e., 10% smoke). As the indication is genetic, we assume that there is no effect of any confounder on the indication (RR: U→Z=1) – just as in Figure 1B. We have limited *a priori* reason to believe that the indication affects lung cancer incidence (beyond that of through the postulated effect of statins). Nonetheless, we will assume that there may exist some weak independent effect (RR: Z→Y=1.30), as certain studies have implicated LDL-C in cancer incidence. Therefore, we have some confounding by indication. Furthermore, we know that smoking has a strong effect on lung cancer (RR: U→Y=10), and we know that smokers are more likely to develop arteriosclerosis and thus receive statins (U→X=1.75). For this hypothetical scenario, there exists no true effect of statins on the outcome (RR: X→Y=1) – as is also supported by recent work on statins and non-small-cell lung cancer [22]. Now, let us take a hypothetical sample of the entire Swedish adult population using nationwide registries (N=8 000 000), from the above specified data-generating mechanism.

Using a modified Poisson regression (model invariant) in an omniscient setting where we have all data available to us (including smoking status), we estimate the least biased RR of statins on lung cancer to 1.00, controlling for smoking status and familial hypercholesterolemia (Table 2). We have bias in our crude estimate (RR 1.39), our familial hypercholesterolemia conditional estimate (RR 1.39), our estimate when we select only individuals without familial hypercholesterolemia (RR 1.38), and our estimate when we select only individuals with familial hypercholesterolemia (RR 2.39) – as we would expect. We have no bias in the estimate where we only control for smoking status, as familial hypercholesterolemia is rare enough to not exert any confounding in that analysis before the second decimal (where Z is not rare, we would expect to see more bias in this analysis). In all analyses using real-world available data (that is, not including smoking status), we would erroneously conclude that statin use has some effect on lung cancer. However, the critical point is that indication-based sampling analysis suffer greater bias than crude analysis (139% vs 39%), despite indication-based sampling breaking the confounding from familial hypercholesterolemia when only studying those with familial hypercholesterolemia (Z=1). This highlights the fact that conditioning on a confounder may result in a net increase in bias. Finally, and of relevance to the applications of epidemiology, indication-based sampling is more biased than a regression adjustment for familial hypercholesterolemia (139% vs 39%), despite the existence of confounding by indication. Indication-based sampling is inferior to regression adjustment regardless of the magnitude of the causal effects considered and the level of confounding – as we will proceed to show using Monte Carlo simulations.

**Table 1.** Tabulation of statin use (X) and lung cancer (Y) over familial hypercholesterolemia (Z) and smoking status (U) from a hypothetical sample of 8 000 000 individuals.

|  | Smoking status=0 |  |  |  | Smoking status=1 |  |  |  |
| --- | --- | --- | --- | --- | --- | --- | --- | --- |
|  | Familial hypercholesterolemia=0 |  | Familial hypercholesterolemia=1 |  | Familial hypercholesterolemia=0 |  | Familial hypercholesterolemia=1 |  |
|  | Cancer=0 | Cancer=1 | Cancer=0 | Cancer=1 | Cancer=0 | Cancer=1 | Cancer=0 | Cancer=1 |
| Statin use=0 | 6 084 471 | 6 120 | 1 8071 | 16 | 580 164 | 5 883 | 528 | 5 |
| Statin use=1 | 1 072 881 | 1 091 | 1 7980 | 20 | 207 164 | 2 098 | 3 470 | 38 |

**Table 2.** The estimated relative risk of statins (X) on lung cancer (Y), depending on whether we control for smoking (U) and familial hypercholesterolemia (Z), either by regression adjustment or indication-based sampling, where the true RR is 1.00.

| Form of analysis | RR | 95% CI | Relative bias |
| --- | --- | --- | --- |
| Regression adjustment for smoking and familial hypercholesterolemia | 1.00 | 0.97-1.04 | 0% |
| Crude analysis | 1.39 | 1.33-1.44 | 39% |
| Regression adjustment for smoking | 1.00 | 0.97-1.04 | 0% |
| Regression adjustment for familial hypercholesterolemia | 1.39 | 1.33-1.44 | 39% |
| Crude among those without familial hypercholesterolemia (indication-based sampling, $Z=0$) | 1.38 | 1.33-1.44 | 38% |
| Crude among those with familial hypercholesterolemia (indication-based sampling, $Z=1$) | 2.39 | 1.45-3.94 | 139% |

### Simulations of bias amplification

Considering that potential bias amplification is dependent on the magnitude of different relationships in a causal system, we performed simulations with different scenarios under the directed acyclic graphs above (Figure 1). Specifically, we perform simulations of scenarios with varying 1) effect of Z on Y, 2) effect of U on Y, and 3) prevalence of Z and U. Scenario one examines the effect of Z on Y as the primary driver of confounding by indication (seeing as Z→X is *a priori* known to be strong), scenario two reflects the weighing of amplification concerns against confounding control of Z, and scenario three examines the impact of sample size as potentially compromised by indication-based sampling and the relevance of Z and U as their influence is minimized at the extremes of prevalence (i.e., 0% prevalence or 100% prevalence).

Except under the scenario where we vary the relevant parameter, we simulate our datasets so that there exists a great effect of the indication on drug use (RR Z→X=10), no effect of the drug on the outcome (RR X→Y=1), no direct effect of the indication on the outcome (RR Z→Y=1), and modest confounding from U on the drug-outcome relationship (RR U→X=1.5 and U→Y=1.5). Except in the simulation where we vary the prevalence’s, we simulate the data so that drug use has a baseline prevalence of 5%, the outcome has a baseline risk of 1%, and both the indication and the confounder have a prevalence of 50%. In favour of simplicity, we assume that there are no other sources of systematic bias than confounding and the arising amplification (e.g., no measurement error or selection bias).

For each scenario, we simulated K=1 000 datasets, each with a sample size of N=1 000 000. In each dataset, we fit a modified Poisson regression. We then obtained the arithmetic mean of the coefficients and 95% confidence interval upper and lower bounds over the datasets. As a measure of intra-scenario variability, we obtained the standard deviation of the coefficients over the K datasets (i.e., the Monte Carlo error on log[RR] scale). To ease interpretability, we exponentiated the obtained mean coefficients to relative risks and their corresponding confidence interval. For a description of each simulation see Appendix.

### Simulation 1: Increasing confounding by indication

Considering that confounding by indication will be greater with an increasing effect of the indication on the outcome, we perform simulations to show how indication-based sampling affects amplifications under a range of effects of Z on Y. Specifically, we allow the RR of Z’s influence on Y to vary between 1 and 2 with increments of 0.10. The Monte Carlo error of the indication-based analysis (least efficient) varied between 0.009 and 0.027 across all scenarios.

We might be inclined to believe that indication-based sampling will be less biased the stronger the effect of Z on Y, as there would be greater confounding by indication. However, the amount of bias in our estimate of X→Y only depends on U in our indication-based sampling analysis (Figure 3). Explicitly, as we have forced X and Z to be independent (by indication-based sampling), the change in Z’s effect on Y does not induce bias in our estimate of X on Y. All remaining bias in our indication-based sample is thus due to U and any amplification – which is also constant across any magnitude of Z→Y.

**Figure 3.**
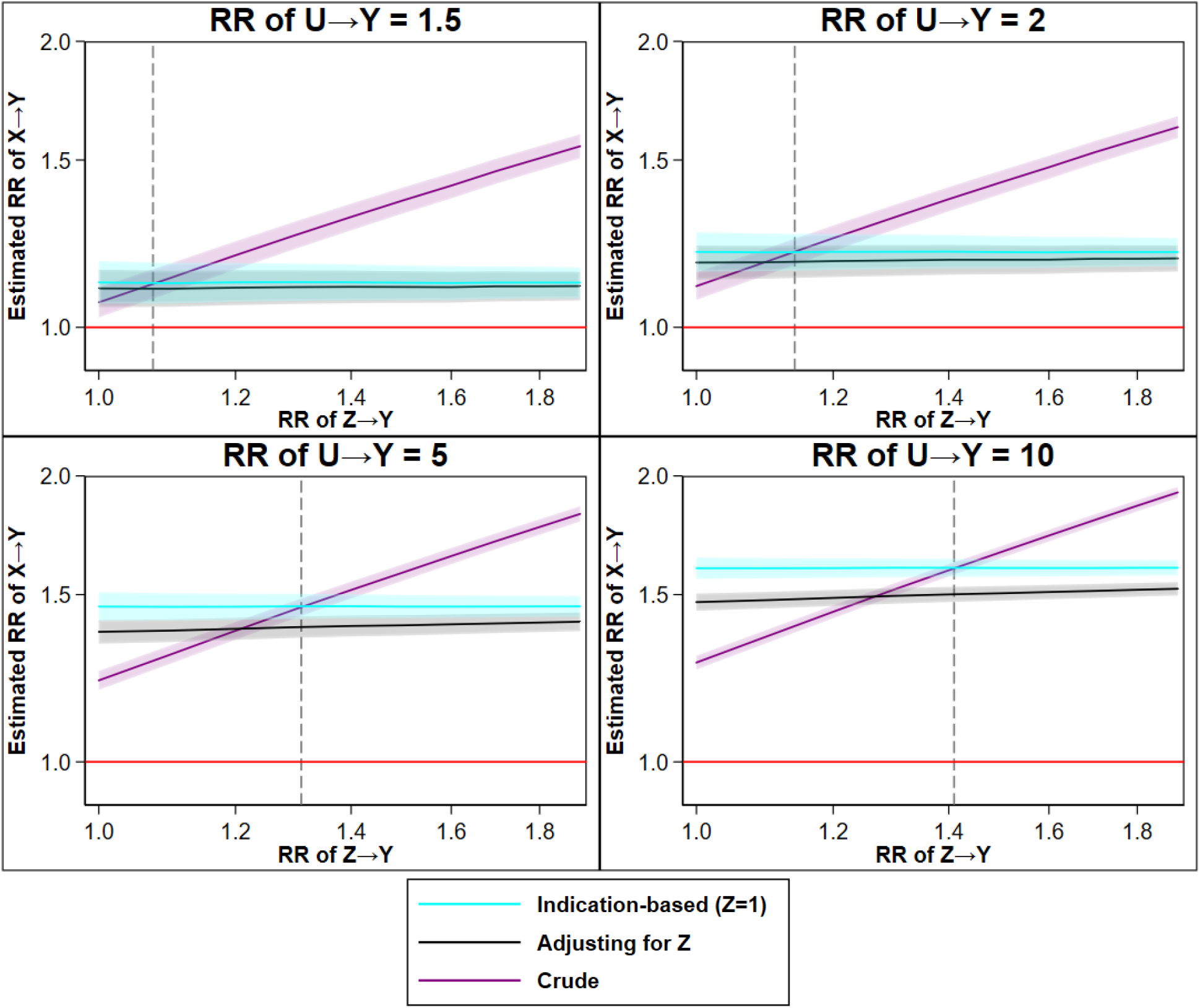
The estimated relative risk of X→Y from a crude analysis (no control), an analysis adjusted for Z (multivariable regression), and an analysis using indication-based sampling (crude analysis while selecting those with Z=1), over ever-increasing confounding by indication (greater Z→Y), by different magnitudes of the effect of the confounder on the outcome (greater U→Y). Legend: Dotted line indicating the preference in terms of net bias between crude and indication-based analysis; on the left side of the dotted line the crude estimator is less biased and on the right side of the dotted line the indication-based analysis less biased. The shaded area indicates a 95% confidence interval. The red line indicates the true causal effect (RR 1). For an extended description of the simulation, see Appendix.

As noted in the hypothetical example of statins and incident lung cancer, indication-based sampling is more biased than standard regression adjustment for Z. This is because indication-based sampling (or other forms of matching) maximizes imbalances in U, while adjusting for Z retains some probability of balances in U between X=1 and X=0 [14]. In other words, indication-based sampling increases the difference in the distribution of U between those with X=1 and X=0, which has implications for the confounding potential of U.

Finally, and as we return to in the next set of simulations, when there exists only minor confounding from U there is minor amplification (RR U→Y≤1.5 under the simulated set-up), and the crude estimate is generally more biased than any method controlling for Z. However, when U has a greater influence (RR U→Y>1.5), the crude estimate may yield a lower net bias as compared to any method controlling for Z (the dotted line in Figure 3 indicates the preference between crude and indication-based analysis).

### Simulation 2: Increasing confounding from the unobserved confounder

As the strength of U will determine the relevance of amplification, we vary the effect of U on Y, in the absence (RR Z→Y=1) and presence (RR Z→Y=1.25) of confounding by indication. Specifically, we vary the RR of U’s influence on Y between 1 and 4 with increments of 0.10. The Monte Carlo error of the indication-based analysis (least efficient) varied between 0.018 and 0.029 across all scenarios.

In the absence of confounding by indication (i.e., when Z is a perfect IV), indication-based sampling and regression adjustment for Z is more biased than crude analysis at all levels of U (Figure 4). This is to be expected given that we have amplification under any method addressing Z, but not under the crude analysis. As such, the crude analysis is only biased by U, while indication-based sampling and regression adjustment are biased by U and amplification of U, yielding a net increase in bias compared to methods not addressing Z. As in examples of increasing Z→Y, indication-based sampling is more biased than standard regression adjustment regardless the effect of U on Y.

**Figure 4.**
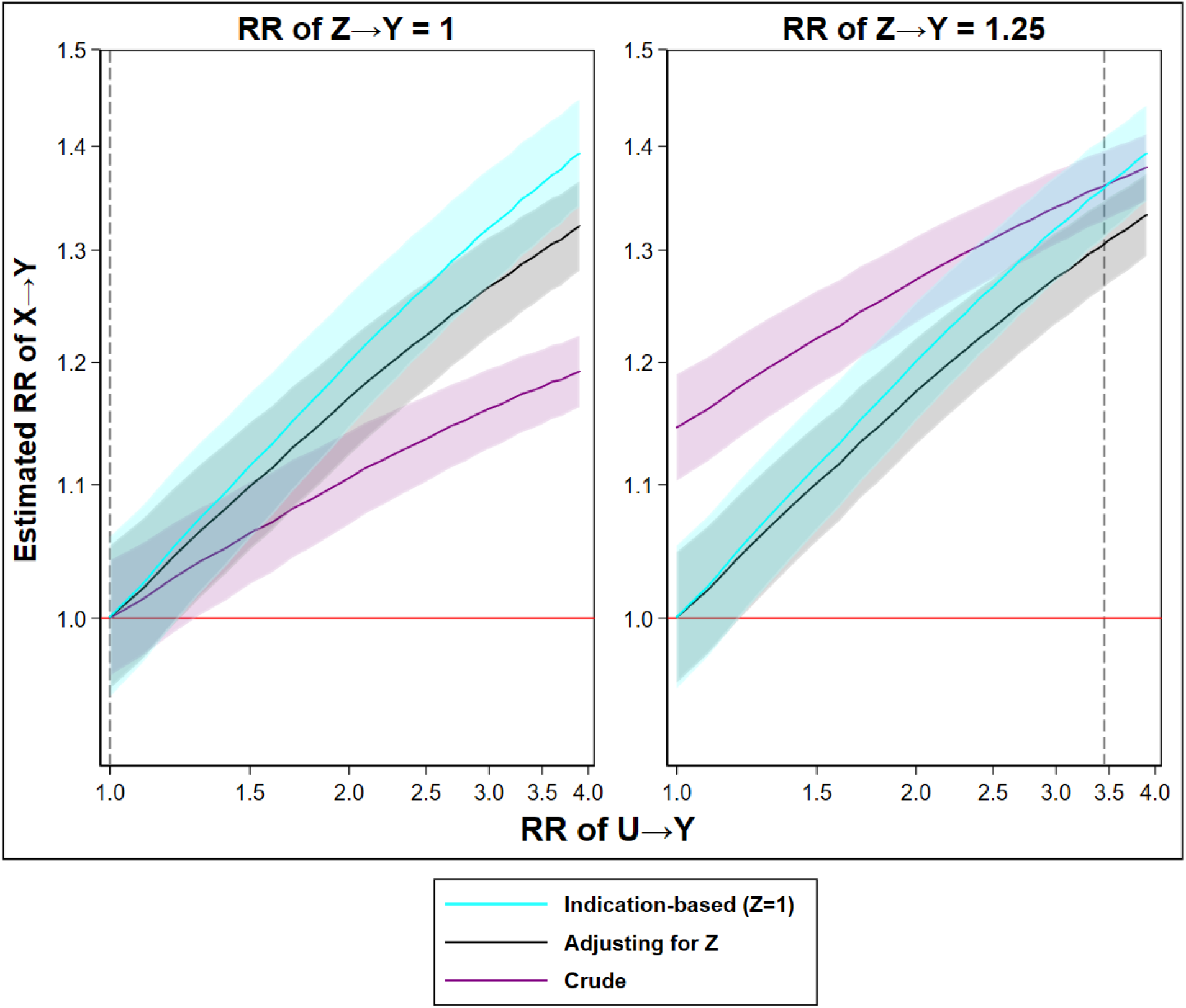
The relative risk of X→Y from a crude analysis (no control), an analysis adjusted for Z (multivariable regression), and an analysis using indication-based sampling (crude analysis while selecting those with Z=1), over ever-increasing confounding from U (greater U→Y), separately whether in the absence or presence of confounding by indication (RR Z→Y: 1.25). Legend: Dotted line indicating the preference in terms of net bias between crude and indication-based analysis. The shaded area indicates a 95% confidence interval. The red line indicates the true causal effect (RR 1). For an extended description of the simulation, see Appendix.

However, in the presence of confounding by indication, the crude analysis may be more biased. Specifically, the crude analysis is biased by both U and Z, while the methods addressing Z (indication-based sampling or regression adjustment) are biased by U and amplification of U. However, any magnitude of U→Y greater than a RR of ∼3.5, under this specific simulation with some confounding by indication, would yield a greater bias in indication-based sampling and regression adjustment than crude analysis. This arises as the benefit of controlling for Z does not outweigh the imposed amplification of U whenever the relative risk of U on Y exceeds 3.5 (under this specific set-up). We stress that this level is fully dependent on the specific set-up (i.e., the data-generating mechanism) and should by no means be considered a generalizable threshold. As in the absence of confounding by indication, indication-based sampling is more biased than standard regression adjustment regardless of the effect of U on Y.

### Simulation 3: Increasing prevalence of the indication and the unobserved confounder

The prevalence of Z and U will influence the optimal analytical strategy, especially as both bias and sample size are concerns for indication-based sampling and pharmacoepidemiology in general. For that reason, we perform our standard simulation with jointly varying levels of prevalence’s of U and Z. The Monte Carlo error of the indication-based analysis (least efficient) varied between 0.017 and 0.089 across all scenarios.

Generally, the greatest amount of bias in the regression adjusted and indication-based approach arise as the prevalence’s of Z and U approach 50%, regardless of whether in the absence or presence of confounding by indication (Figure 5). As always, in the presence of confounding by indication, the crude estimator is generally more biased than the alternatives. At the extreme tails of the prevalence distributions, all estimates will approximate the true causal effect. This occurs as the variability in Z and U decreases, as either all individuals have Z and U or no one does – resulting in exchangeability over X. In the latter extreme, it is of course impossible to perform Z=1, as no one fulfils that condition.

**Figure 5.**
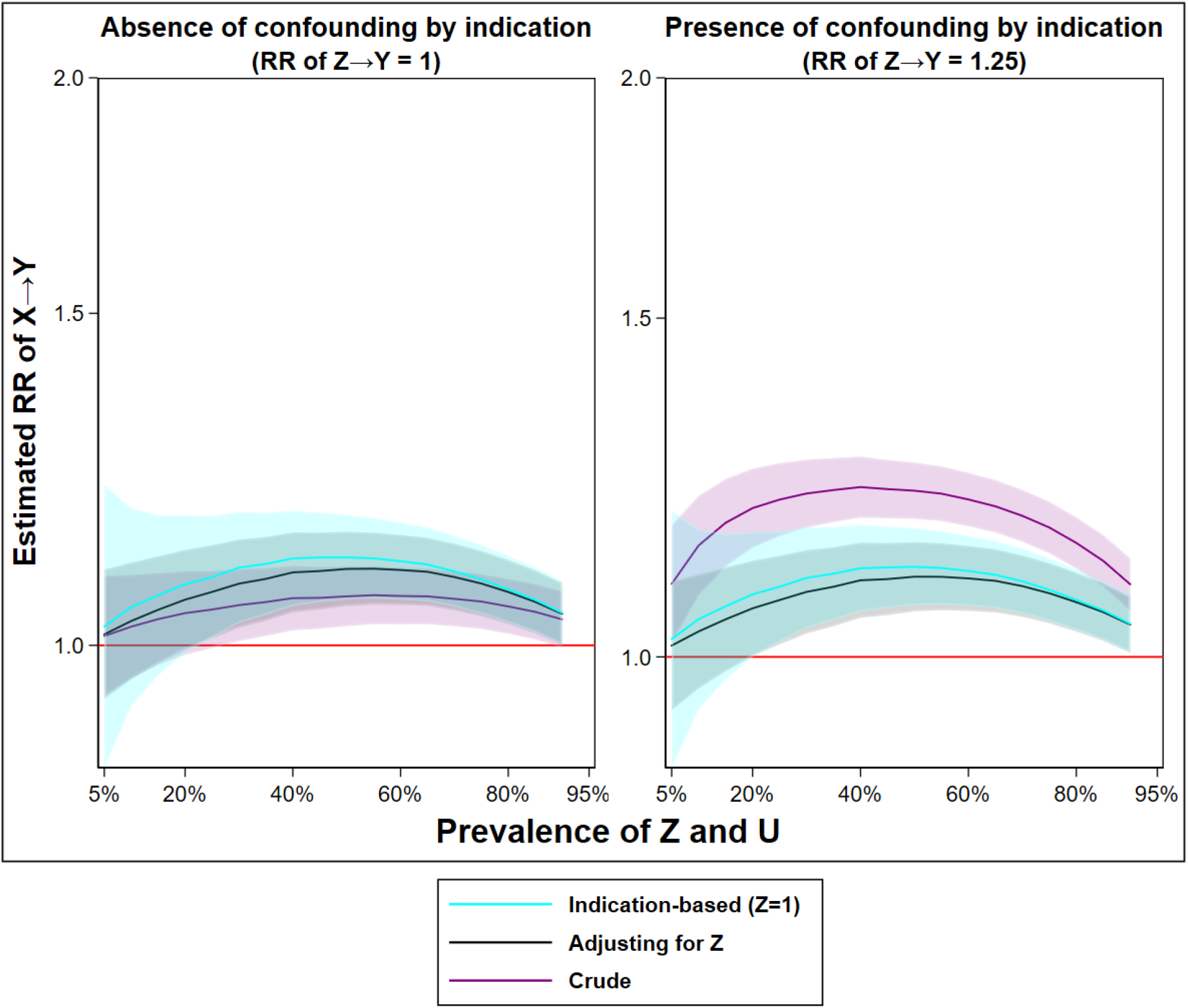
The estimated relative risk of X→Y from a crude analysis (no control), an analysis adjusted for Z (multivariable regression), and an analysis using indication-based sampling (crude analysis while selecting those with Z=1), over an ever-increasing prevalence of Z and U and by whether confounding by indication is absent or present (RR of Z→Y = 1.25). Legend: The shaded area indicates a 95% confidence interval. The red line indicates the true causal effect (RR 1). For an extended description of the simulation, see Appendix.

However, indication-based sampling can suffer additional substantial bias when the prevalence of U and Z is low. This is because there is a finite number of individuals satisfying the condition Z=1, which could result in an inflation of chance imbalances, amplification magnitude, and random error when the prevalence is low. Specifically, as we restrict to Z=1 and as the prevalence of Z decreases, we reduce the sample and leave the analyses strongly influenced by fluctuations in all bias parameters (including random noise). The random error component of amplification in finite samples is distinct from the systematic component of bias amplification which has an expected direction and magnitude given knowledge about the true relationships between factors in a causal system (i.e., the data-generating mechanism). We chose to emphasise this scenario as it has implications for indication-based sampling performed in pharmacoepidemiology of indications with relatively low population prevalence (e.g., epilepsy). Notably, the Z regression adjustment model does not suffer the same volatility as indication-based sampling, although the model will fail to estimate the Z coefficient in circumstances where Z is extremely rare (and positivity could be violated).

### Drug effect-heterogeneity

We have yet to consider drug effect-heterogeneity, which implies that there exist distinct clinically relevant effects among certain patient groups, often defined by the indication. Specifically, if there exists true heterogeneity in drug effects depending on the indication (i.e., effect-modification) it may be warranted to perform indication-based sampling. For example, it is only relevant to study the effect of antidepressants on suicide attempts among the patient population who experience an indication (e.g., severe depression), since it is unlikely that those without indication have any benefit of the therapy. Yet, it is still true that this will lead to bias amplification in the presence of an unobserved confounder (as shown above). Such amplification is, however, likely worth the trade-off since effect-heterogeneity will outweigh the induced bias.

However, in many pharmacoepidemiological studies, there is no *priori* reason to believe that effect-heterogeneity exists. For example, a common application of pharmacoepidemiology is the pharmacovigilance of drug teratogenicity, since clinical trials rarely include pregnant populations. For instance, many studies have examined the relationship between certain antiseizure medications (especially valproic acid) and congenital malformations [24]. There is no evidence that antiseizure medications teratogenicity is unique to any indication. Rather, the teratogenicity is believed to result from a disruption of embryological development that is constant across pregnancies. In such a scenario, there is no immediate benefit to employing indication-based sampling, even to overcome confounding by indication. Specifically, in the absence of effect-heterogeneity, the optimal strategy to overcome confounding is to perform some adjustment (e.g., regression methods), and not to perform indication-based sampling as this will maximize bias amplification and drastically reduce the eligible sample.

In scenarios where there exists no true effect-heterogeneity and we erroneously employ indication-based sampling, we are in fact likely to mistake bias amplification for heterogeneity. Consider the simple dramatic example (Table 2), where we estimate the relative risk to 1.38 (1.33-1.44) when forcing Z=0, and to 2.39 (1.45-3.94) when forcing Z=1. Were we to perform an arbitrary statistical test, we might conclude that these are different, and may attribute the difference to effect-heterogeneity. This is an erroneous conclusion, which we know since we designed the simulation so that it does not include effect-heterogeneity – we are merely detecting bias amplification. This erroneous conclusion has been noted in passing in prior examinations of bias amplification (i.e., Appendix 1 in Myers et al. [18]). Unfortunately, there is no way to distinguish amplification and effect-heterogeneity using observed data, and we must rely on expert knowledge about the world.

For the sake of completeness, we wish to note that it is possible to overcome this trade-off between effect-heterogeneity and amplification by assuming that Z does not affect X. Specifically, if Z has no causal effect on X then Z stratification will not lead to bias amplification, although such analysis will still be influenced by any confounding from other factors. Nevertheless, we believe that this “independence” assumption is of limited practical relevance in pharmacoepidemiology, where Z (indication) always has a strong causal effect on X (drug use).

In summary, while a total risk-benefit assessment may need to consider stratum-specific effects under effect-heterogeneity for specific outcomes (e.g., depression in the study of antidepressants and suicide), it can be detrimental to perform indication-based sampling in the study of outcomes where there is no *a priori* reason to believe effect-heterogeneity exists (e.g., the study of antiseizure medication teratogenicity).

## Discussion

We have provided, to the best of our knowledge, the first description of how bias amplification arises under indication-based sampling and outlined how this problem applies in pharmacoepidemiology. While it is true that indication-based sampling alleviates confounding from indication, we have highlighted that it may result in bias amplification, a self-inflicted injury that can result in a net bias increase from unobserved (or residual) confounding. As such, if unobserved confounding is the scary monster in the corner of the room of epidemiology, then bias amplification is the equivalent of giving that monster a baseball bat. We have further shown that indication-based sampling is more biased than alternative approaches to control for drug indication (e.g., regression adjustment). This suggests that even if confounding by indication exists, it is often not warranted to perform indication-based sampling, an approach that also depletes the available sample size. Finally, we have detailed how bias amplification may be erroneously interpreted as effect-modification, while acknowledging that the need for assessment of true effect modification may sometimes outweigh concerns of bias amplification. As such, we suggest that bias amplification should be considered in all aetiological studies using observational data and should be specifically addressed if indication-based sampling is performed.

Importantly, bias amplification also arises if one selects a cohort without indication (Z=0 stratification), and the magnitude and direction depend on the relationships between factors. Specifically, the same concerns apply if one selects a cohort without indication (Z=0) as those described for indication-based sampling (Z=1), and as such, neither the presence nor absence of indication are ideal eligibility criteria - despite previous authors suggesting that restriction to those without clinically manifested indication may be a good strategy to overcome confounding by indication [25].

While the expected value of the systematic bias originating from amplification can be outlined in an omniscient setting with an infinite sample, such expectations are unlikely to be true in finite samples. Specifically, since the eligible sample size is often substantially compromised by indication-based sampling, the random error component of bias amplification is likely to yield an unexpected magnitude and direction in a single study. To reiterate, while we can expect how the systematic bias from bias amplification will influence our estimates if we had an infinite sample and if we were all-knowing, we have little reassurance that this expected bias direction and magnitude will be true in the empirical setting.

While our focus has been that of traditional pharmacoepidemiology, our reasoning extends to any investigation using observational data. For example, bias amplification has already been recognized in the propensity score literature [12, 13], where the inclusion of instrumental variables (Z) in a propensity score in the presence of unmeasured confounding leads to bias amplification (sometimes referred to as Z-bias [15, 20]). Furthermore, bias amplification is the same phenomenon that leads to bias in within-family/trio mendelian randomization when one parent’s genotypes are omitted [26]. It is also bias amplification that gives rise to inflation of non-shared confounding in sibling analyses [16, 27].

We further note that reasoning about bias amplification is largely incompatible with the philosophy of the emulated target-trial framework [4, 5], as such a framework assumes conditional exchangeability and justifies indication-based sampling on the premise that only those with indication would be enrolled in a hypothetical trial. We argue, as others before us [18], that bias amplification should be carefully considered when reasoning about the observational emulation of a target trial and that restriction on indication should be avoided if possible in the data collection phase.

### Limitations

Despite our efforts to be thorough in our descriptions, there are limitations of our work that must be acknowledged. First, as with any simulation study, there is an abundance of scenarios that we have not considered nor explored. The relationships between factors (data-generating mechanisms) that we have outlined are simplifications of reality and constitute just some of an infinite number of possible mechanisms which could have been considered. One specific scenario which we have not considered is that in which the unobserved confounder (U) has some non-negligible effect on indication (Z). While such a scenario could, sometimes, justify control for indication even if it was a true IV (i.e. indication has no effect on the outcome) – to alleviate part of the imposed confounding by U – its implications must be considered in the light of recent work on trapdoor variables [28]. Nonetheless, we have favoured simplicity to enhance transportability outside our considered mechanisms.

Second, we recognize that we have relied on informal epidemiological definitions of confounders. Unfortunately, even if one were to consider formal definitions of confounders, as those under the counterfactual framework [29], such definitions provide no utility when reasoning outside the omniscient setting as they rely on information about the true bias parameter. Yet, we note that were one to define confounders according to such formalizations one would be able to make the distinction between a bias amplifier and confounder based on the net effect on bias after controlling for their influence.

Finally, we have not considered a scenario where there exist multiple forms of bias. For example, we have not considered typical epidemiological bias processes such as misclassification. Although bias amplification may be considered a systematic error that will generally be independent of other processes, it should be noted that if there, for example, exists misclassification of the indication (Z) then the observed amplification will be distinct from the true amplification (i.e., in the absence of misclassification of Z). Future work may be directed at studying the complex interplay between multiple bias processes in the presence of residual confounding, and especially recognizing the random error component of bias processes in epidemiology.

### Practical implications

While further research is needed on bias amplification, especially considering more complex scenarios than those generated here, we believe that there is some immediate practical utility of our work. Specifically, current risk-benefit analyses used by national agencies are partly based on observational analyses using indication-based sampling. For example, health technology assessments often leverage indication-based sampling when evaluating post-market effectiveness and safety, or agencies may require the employment of indication-based sampling by pharmaceutical companies when such companies are to estimate the long-term efficacy for subsidy decisions. While the employment of indication-based sampling may be warranted in scenarios where there exists true effect-modification, such analytical choices should be scrutinized to identify possible bias amplification.

Unfortunately, identifying the most appropriate analytical decision requires substantive knowledge about the true relationships between different factors (i.e., the data-generating mechanism). While there exist principles that should guide covariate control, such as the preference of factors associated with the outcome of interest [17, 18], it is more difficult to distinguish bias amplification from effect-modification. Assessing the consistency of associations across analytical approaches (i.e., analytical triangulation) is, however, one feasible strategy if data availability allows. Similar estimates of associations from indication-based sampling and regression adjustment analysis either indicate that all bias processes perfectly balance, or that no bias amplification and no effect-modification exist. Principles of Occam’s razor suggests that the latter is more probable. It is, however, not possible to infer the cause of an eventual inconsistency between analytical approaches since it can be either because of amplification or effect-modification. As such, this analytical triangulation approach may serve as a rule-out check. Nevertheless, this approach may be readily employed in settings where data availability allows investigators to use a multitude of analyses (e.g., in studies using electronic health/medical records data). Unfortunately, and perhaps obviously, it is not possible to perform such checks if a clinical cohort has been enrolled using indication-based sampling, such as in pharmacovigilance studies of drug teratogenicity [10]. The challenge of bias amplification arising in clinical cohorts enrolling based on indication makes us inclined to re-state the take-home message of our work: bias amplification should be considered in the list of biases and other considerations during the design phase of any pharmacoepidemiological investigation using observational data.

## Conclusion

Studies using indication-based sampling should have robust justification. This approach should by no means be considered a “free-lunch” as indication-based sampling is generally more biased than alternative modes of analysis. As such, we suggest that future observational studies stay wary of bias amplification when considering drug indications.

## Data Availability

All data produced in the present work are contained in the manuscript

## Acknowledgement

The study was funded by the National Institute of Health (1R01NS107607-01A1), Erik and Edith Fernström Foundation for Medical Research (2020-00321), Karolinska Institutet (2020-00160, 2020-01172) and the Swedish Society for Medical Research (RM21-0005). The funders had no role in the design of the study, data management, data analysis, interpretation of findings and the decision to submit the article for publication. This work was carried out using the computational facilities of the Advanced Computing Research Centre, University of Bristol - http://www.bris.ac.uk/acrc/.

