## Appendix for "Bias amplification of unobserved confounding in pharmacoepidemiological studies using indication-based sampling: there is no free lunch in restricting the sample to those with a particular drug-indication"

### Simulation specifications

#### The data-generating mechanism for the simple example

For each individual:

$P\left( U=1 \right)=.1$,

$P\left( Z=1 \right)=.005$,

$P\left( X=1 | U,Z \right)=\alpha_{0}\alpha_{1}^{U}\alpha_{2}^{Z}=.15*{1.75}^{U}*{3.33}^{Z}$,

$P\left( Y=1 | U,X,Z \right)=\beta_{0}\beta_{1}^{U}\beta_{2}^{X}\beta_{3}^{Z}=.001*{10}^{U}*1^{X}{*1.30}^{Z}$,

#### Varying Z→X

For each individual:

$P\left( U=1 \right)=.5$,

$P\left( Z=1 \right)=.5$,

$P\left( X=1 | U,Z \right)=\alpha_{0}\alpha_{1}^{U}\alpha_{2}^{Z}=.05*{1.5}^{U}*{10}^{Z}$,

$P\left( Y=1 | U,X,Z \right)=\beta_{0}\beta_{1}^{U}\beta_{2}^{X}\beta_{3}^{Z}=.01*{1.5}^{U}*1^{X}{*\tau}^{Z}$,

Where we allow Z’s influence on Y to be equal to τ. We vary τ between 1 and 2 with increments of 0.10. For each scenario, we simulated 1000 datasets with N=1 000 000. In each dataset, we fit a modified Poisson regression (i.e., Poisson regression with a robust error variance). We then average the obtained coefficients and their bias over the datasets at each τ.

#### Varying prevalence of Z and U

For each individual:

$P\left( U=1 \right)=\tau$,

$P\left( Z=1 \right)=\tau$,

$P\left( X=1 | U,Z \right)=\alpha_{0}\alpha_{1}^{U}\alpha_{2}^{Z}=.05*{1.5}^{U}*{10}^{Z}$,

$P\left( Y=1 | U,X,Z \right)=\beta_{0}\beta_{1}^{U}\beta_{2}^{X}\beta_{3}^{Z}=.01*{1.5}^{U}*1^{X}{*1.25}^{Z}$,

We vary τ between 0.05 and 0.95 with increments of 0.05. For each scenario, we simulated 1000 datasets with N=1 000 000. In each dataset, we fit a modified Poisson regression (i.e., Poisson regression with a robust error variance). We then average the obtained coefficients and their bias over the datasets at each τ. For the absence of confounding by indication we fix $\beta_{3}^{Z}=1$ (instead of 1.25).

#### Varying U→Y

For each individual:

$P\left( U=1 \right)=.5$,

$P\left( Z=1 \right)=.5$,

$P\left( X=1 | U,Z \right)=\alpha_{0}\alpha_{1}^{U}\alpha_{2}^{Z}=.05*{1.5}^{U}*{10}^{Z}$,

$P\left( Y=1 | U,X,Z \right)=\beta_{0}\beta_{1}^{U}\beta_{2}^{X}\beta_{3}^{Z}=.01*\tau^{U}*1^{X}{*1.25}^{Z}$,

Where we allow U’s influence on Y to be equal to τ. We vary τ between 1 and 4 with increments of 0.10. For each scenario, we simulated 1000 datasets with N=1 000 000. In each dataset, we fit a modified Poisson regression (i.e., Poisson regression with a robust error variance). We then average the obtained coefficients and their bias over the datasets at each τ. For the absence of confounding by indication we fix $\beta_{3}^{Z}=1$ (instead of 1.25).
